# The First-Hand Needs of Informal Caregivers of People Living with Dementia, in Residential Care Settings: A Scoping Review

**DOI:** 10.1101/2025.01.18.25320759

**Authors:** Chloe Moody, Jeremy Dixon

**Affiliations:** Centre for Death and Society, University of Bath; Centre for Adult Social Care Research (CARE), Cardiff University

## Abstract

**Background:** Dementia is a terminal condition often requiring palliative care delivered in residential care settings. While informal caregivers (ICGs) are pivotal in care-based decision-making, they have higher rates of physical and mental illness than ICGs of people with other terminal conditions. Identifying the needs of ICGs of people living with dementia (PLwD) is essential, to mitigate these risks and develop effective support systems.

**Objective:** Our objective was to understand the first-hand experiences and needs of ICGs of PLwD receiving palliative and end-of-life care, in residential care settings.

**Method:** Following the JBI methodology for scoping reviews, electronic databases (APA PsychNet, the Cochrane Database of Systematic Reviews, PubMed and Web of Science) were searched in September 2024, with no publication date limitations. Thematic synthesis was conducted on the findings of eligible peer-reviewed and grey literature, written in English, and reported in accordance with the PRISMA-ScR checklist.

**Results:** Forty-six articles were included. There were three overarching themes: “knowledge and understanding of dementia”, “engagement in care-based decisions”, and “coping mechanisms and support for own wellbeing”. Sub-themes presented an interplay between these, demonstrating the importance of understanding dementia, the significance of such knowledge for ICGs to maintain their own wellbeing, subsequently influencing their engagement in care-based decision making.

**Conclusion:** Care settings must work towards compassionate and timely support for ICGs, including a stable point of contact throughout admission and should use lay language. Future studies should take a longitudinal approach to understand the evolving role of ICGs, with particular attention to cultural and ethnic needs.

**What is already known on this topic:**

- Alongside care-staff, informal caregivers play a critical role in care-based decision-making and support for people living with dementia (PLwD), but they face significant health and wellbeing challenges, with limited research on effective mechanisms to involve and support them in their role.

**What this study adds:**

- Our study highlights the interconnected challenges faced by informal caregivers of PLwD in understanding dementia, maintaining their own wellbeing, and engaging in care-based decisions for PLwD.

**How this study might affect research, practice, or policy:**

- The findings identify a need for proactive, compassionate support for informal caregivers of PLwD through navigable resources, education surrounding dementia terminology using lay-language, and consistent communication with care-staff to build trust and stability for informal caregivers.
- Our scoping review highlights the need for longitudinal research on evolving informal caregiving roles and calls for further research to explore and address the diverse needs of underserved communities, to inform culturally competent policies and practices.

## INTRODUCTION

### Rationale

‘Dementia’ is an umbrella term for conditions which lead to cognitive impairment and decline of brain functioning, such as Alzheimer’s disease, Lewy bodies, and vascular dementia (1). A life-limiting and progressive condition, dementia was ranked the seventh leading cause of death in 2021 globally, accounting for the deaths of 1.8 million people (2). Towards end of life, people living with dementia (PLwD) may require assistance to maintain their health and wellbeing, which can be delivered through palliative or end-of-life care services (3, 4). International comparative research indicates that between 17.5% and 77.3% of PLwD die in residential care settings (such as care homes or nursing homes), indicating the need to focus on palliative care provision within these services (5).

Whilst much of the day-to-day care for PLwD in residential care settings will be provided by professional carers, the role of informal caregivers remains significant. Informal caregivers are individuals who maintain a meaningful relationship with PLwD, whilst providing unpaid support to ensure their daily health and wellbeing (6, 7). They may be family members or may have a non-familial relationship with the person, such as being a close friend. Informal caregivers have a significant role in the care of PLwD within residential care settings including decision-making relating to their admission to the home (8), advanced care planning (9), and surrogate decision-making (10).

Policymakers have indicated that health and care services should be aware of the needs of the informal caregivers of PLwD; supporting them to ensure their health and wellbeing whilst involving them in care-based decision making processes (11, 12). However, current research shows significant gaps in each of these areas. Research indicates that informal caregivers’ needs are diverse, and though they often wish to preserve feelings of interconnectedness with the PLwD, they might require support from the residential care setting with structuring visits to enable them to share meaningful moments with that person (13). Some authors argue that research focussing on what might facilitate better family involvement within long-term care settings remains limited, with few studies providing evidence as to the mechanisms or care components which might engender this (14). Furthermore, it is unclear what supports might be offered to carers of PLwD in residential care settings to improve their own health and wellbeing. Existing literature repeatedly demonstrates the negative impact of caregiving on informal caregiver health and wellbeing, often restricting their ability to care for themselves (15, 16). Moreover, family caregivers of PLwD have been highlighted as particularly vulnerable to depression, anxiety, and poorer physical health, in comparison to informal caregivers of those with other terminal conditions (17). Subsequently, there is a need for health and social care professionals to understand the experiences of informal caregivers of PLwD in residential care settings, so risks to their health and wellbeing can be minimised and effective service developed. A preliminary search of the Cochrane Database of Systematic Reviews and JBI Evidence Synthesis was conducted and indicated that no current or ongoing systematic reviews on the topic were in progress, indicating a significant gap in the literature.

### Objectives

This review’s objective is to understand the extent of literature surrounding first-hand experiences of informal caregivers of PLwD receiving palliative or end-of-life care in residential care settings. This review aims to identify themes within the identified ‘needs’ of informal caregivers, understanding how they would like to be involved and supported, and acknowledge any gaps within the literature.

Our primary research question was, ‘What is known about the first-hand experiences and needs of informal caregivers of PLwD receiving palliative & end-of-life care, in residential care settings?’. Within our review, the following sub-questions were used to understand this phenomenon:

1. What are the ‘needs’ of informal caregivers who have cared for a person living with dementia, receiving palliative or end-of-life care, in residential care settings?
2. How would informal caregivers like to be involved in the process of PLwD receiving palliative or end-of-life care in residential care settings?
3. How would informal caregivers like to be supported to promote their own health and wellbeing?

## METHODS

### Protocol and registration

To gain a thorough understanding of the first-hand experiences and needs of informal caregivers of PLwD in this context, this scoping review collated all peer-reviewed and grey literature exploring this topic (qualitative, quantitative, and mixed-method approaches). A protocol was produced prior to conducting this review, in adherence to the JBI methodology for scoping reviews, and is available to access on the Open Science Framework (https://osf.io/vr2hu)(18, 19). Furthermore, this paper has been written according to the Preferred Reporting Items for Systematic Reviews and Meta-Analyses extension for Scoping Reviews (PRISMA-ScR) (19).

### Eligibility criteria

Peer-reviewed studies and grey literature, such as dissertations, written in the English language were eligible for inclusion. There were no limitations on publication date to provide a comprehensive review of the literature. The first search was conducted in August 2023, and a second search was completed in October 2024 to update the paper searches.

Eligible literature needed to include participants who identified as an informal caregiver of PLwD who had permanently transitioned into a residential care setting, and was receiving palliative or end-of-life care. For this review, informal caregivers were defined as unpaid carers that provided care outside of a professional capacity which included, but was not limited to, family or friends. Additionally, residential care settings were considered as settings where PLwD have moved into to receive care outside of their own home, where the responsibility of caregiving no longer solely relied on themselves, or an informal caregiver. For inclusion, eligible studies needed to record the first-hand perspectives of informal caregivers, but studies that reported these perspectives alongside the views of other stakeholders such as professional caregivers, were also eligible for inclusion.

### Information sources and search strategy

Searches were conducted on APA PsychNet, the Cochrane Database of Systematic Reviews, PubMed and Web of Science, using the Boolean search: ((“dementia” OR “people living with dementia” OR “PwD” OR “lewy body dementia” OR “lewy bodies” OR “Alzheimer’s”) AND (“hospice” OR “care home” OR “nursing home” OR “residential care setting” OR “residential care”) AND (“self-reported” OR “interview” OR “lived experience” OR “expectations” OR “needs” OR “experiences” OR “support” OR “preferences”) AND (“hospice care” OR “palliative care” OR “palliative” OR “end of life”) AND (“family caregiver” OR “family” OR “loved ones” OR “primary caregiver” OR “caregiver” OR “informal caregiver”)).

### Selection of sources of evidence

All citations, retrieved from the paper searches, were saved using Endnote Online Classic (Clarivate Analytics, PA, USA). Upon removing all duplicates, two independent reviewers (CM and JD) screened papers according to their titles and abstracts for inclusion. To test the compatibility in reviewer screening and set a mutual understanding on the parameters of study eligibility, the reviewers independently reviewed the full-texts of the first ten papers and discussed their decisions and justifications for paper inclusion and exclusion. The full-texts of the remaining papers were then screened independently, with the two independent researchers (CM and JD) producing lists of studies they decided to include and exclude. Upon completion of full-text screenings, both reviewers compared their lists to identify any discrepancies. Any discrepancies were then discussed and resolved, to provide a final list of studies eligible for inclusion and thematic analysis. This process was then repeated in October 2024, when the updated search was conducted.

### Data charting process

Findings from eligible studies were manually extracted into an adaptation of the JBI extraction tool (18). Digressing from our initial data extraction tool, ‘Study context’, ‘method’ and ‘identified support or coping strategies’ were removed, and ‘Coping mechanisms or support mentioned to improve own health and wellbeing’ were added. These changes were made to refine the data collection and to make it more relevant to the research questions of this scoping review.

### Data items

The final data extraction tool subsisted of the following:

- Year of publication
- Citation
- Location of study
- Research Questions/Aims
- Sample (relationship to PLwD)
- Identified needs of informal caregivers
- Coping mechanisms or support mentioned to improve own health and wellbeing
- Preferences and/or suggestions for how informal caregivers could be involved in the decisions and care of PLwD
- Limitations
- Implications for practice
- Avenues for future research

### Synthesis of results

A thematic synthesis was conducted on findings to provide ‘descriptive’ and ‘analytic’ themes (20). We were unable to provide a meta-analysis of quantitative data as this data was heterogenous, so data was produced in a narrative form. Themes surrounding the practical needs, desired support and coping mechanisms for informal caregivers’ own wellbeing, and preferences on care involvement were produced to provide an initial framework for data charting. Supporting figures and tables were expanded upon through narrative summaries, directly linking the findings to the research questions of this scoping review.

## RESULTS

### Selection of sources of evidence

See figure 1 for a flow diagram of the complete paper screening process.

**Figure 1.**
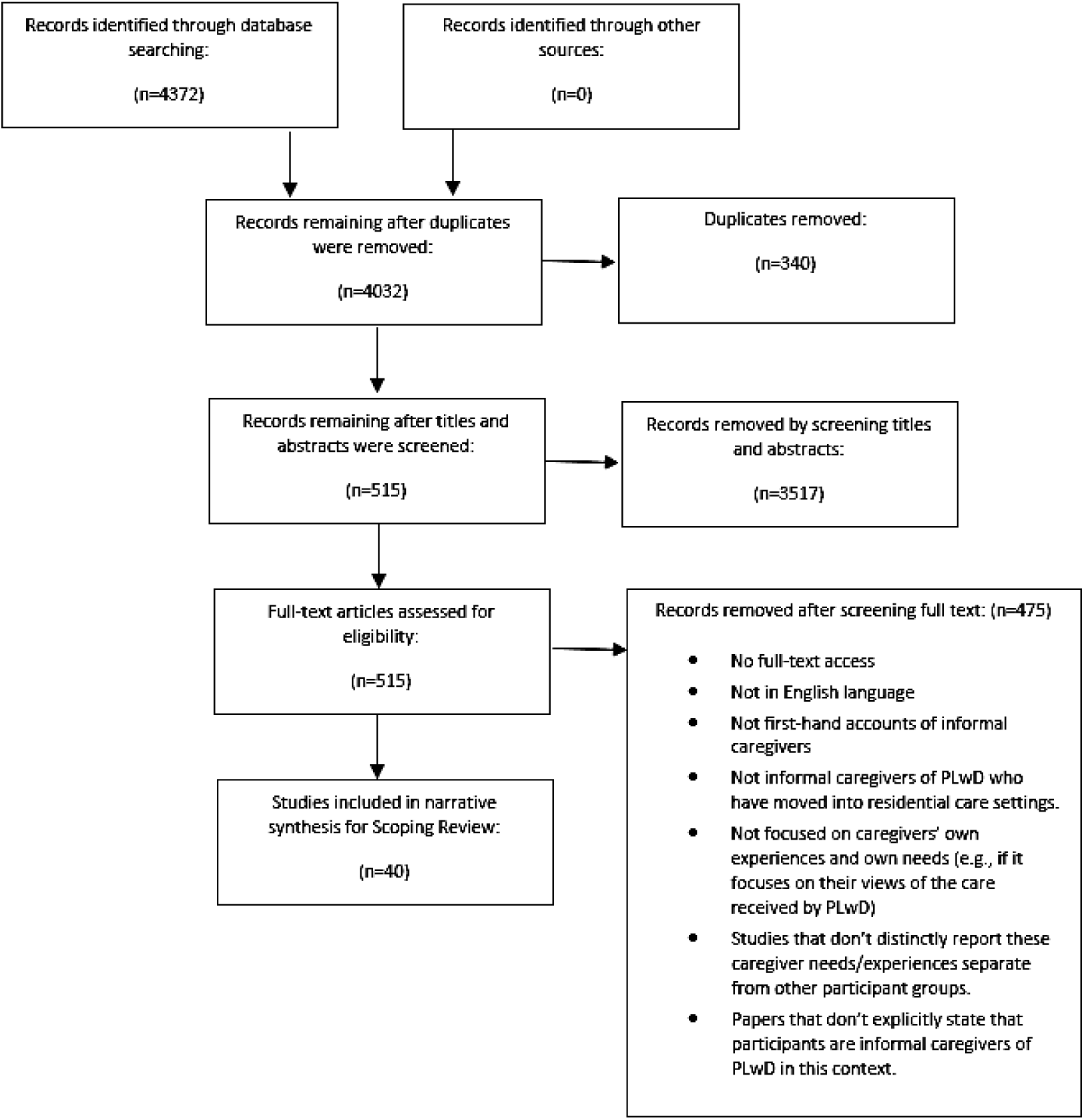
Flow diagram showing the process of paper screening, in accordance with this scoping review’s inclusion criteria.

### Characteristics and Results of Sources of Evidence

Forty-six eligible articles were included for review, all produced between 2000 – 2024. Quantitative studies (n=2) (21, 22), qualitative studies (n=39)(23–61) and mixed methods studies (n=4)(62–65) were included, with only one thesis (24) constituting the grey literature contributing to this review’s findings. The majority of included studies have been conducted in Western countries, including the USA (n=13)(23, 26, 33–35, 39, 42, 43, 45, 49, 52, 57, 59), UK (n=11)(24, 25, 28, 30, 37, 50, 51, 61–64), Canada (n=4)(27, 31, 32, 56), Australia (n=2)(29, 55), Netherlands (n=2)(40, 41), Norway (n=1)(47), Spain (n=1) (53) and Sweden (n=1)(54). Hennings et al (37) conducted their study across North America, UK, and Australia, and McLennon et al (46) conducted their study in the UK and USA. Brazil et al (58) conducted their study across nursing homes in Canada, Czech Republic, Italy, Netherlands, Republic of Ireland, and the UK. One study had been conducted in Israel (21), one in Italy (60), and four articles were reviews, and therefore did not have a specific location of data collection (36, 44, 48, 65).

Predominantly, articles utilised the term ‘family caregiver’ (n=21) when describing their participant sample. Some papers, however, specified the relationship between PLwD and informal caregivers. Please see Supplementary Table 1 for a detailed overview of these identified relationships to participants.

### Synthesis of results

Please see Supplementary Table 1 for a comprehensive presentation of the data extracted from each source of evidence, in accordance with the scoping review objectives and research questions.

## DISCUSSION

Three overarching themes were identified: “knowledge and understanding of dementia”, “engagement in care-based decisions”, and “coping mechanisms and support for own wellbeing”. Emergent sub-themes presented an interplay between these three themes, demonstrating the importance of understanding dementia, how this played a significant role in the coping of informal caregivers’ to maintain their own wellbeing, and subsequently mediate their engagement in care-based decisions for PLwD.

### Theme 1: Knowledge and understanding of dementia

Theme 1 refers to informal caregivers’ fundamental need for a knowledge and understanding of dementia. Education and awareness of the cared-for person’s condition was perceived as key to their engagement in care-based decisions. Such knowledge was also seen as important to maintaining their own wellbeing. The literature described a wide variation of provisions offered by residential care settings, to educate informal caregivers about dementia. Particularly highlighting support which informal caregivers, irrespective of their decision to engage in their role, have acknowledged as key to easing their experience.

#### Setting expectations

Sixteen articles stressed the importance of receiving a clear diagnosis from care professionals, with informal caregivers stating that this enabled them to have realistic expectations about the trajectory of their person’s condition (see Supplementary Table 1). Participants particularly iterated the importance of understanding the prognosis and trajectory of the type and severity of PLwD’s dementia, in the early stages of their diagnosis (23, 24, 27, 29, 31, 33, 39, 42, 45, 50, 52, 54, 56, 57, 63, 65).

Three studies identified that informal caregivers felt that accepting dementia as a terminal condition was less detrimental to their wellbeing, as opposed to facing the uncertainty regarding the trajectory of a person with dementia (23, 33, 65). Illustrated by a participant quote describing, “A crazy roller coaster at the end”, Durepos et al (32) emphasised the importance of setting realistic expectations of care trajectories for informal caregivers through three key concepts. These were; gaining a sense of control within their situation, fulfilling their obligations as a caregiver, and adapting to the loss of PLwD. In a similar vein, Durepos et al (32) showcased the experiences of bereaved informal caregivers of PLwD to demonstrate the strain that a lack of preparedness might cause, both physically and emotionally.

Informal caregivers disliked optimistic statements giving false hope or misrepresentations of the reality of dementia (42), with some demonstrating the need to witness the deterioration of PLwD’s condition first-hand (32, 39). Black et al (26) highlighted the benefits of informal caregivers witnessing the decline of other care home residents living with dementia, and the experiences of their informal caregivers, as a means through which they might prepare themselves for similar experiences.

#### Guidance and signposting

Twenty-four studies highlighted a need for, and challenges associated with, sourcing information and support services (see Supplementary Table 1). Often represented through themes of challenges and barriers to access, informal caregivers underscored the need for organisations and care settings to provide information, guidance, and to have navigable access to support services.

Though three studies stressed the need for services to educate informal caregivers on key terminology surrounding dementia, end-of-life, and palliative care (42, 51, 52), five articles expressed a need for care-staff and services to utilise lay-language when delivering information surrounding PLwD’s condition and care (31, 36, 45, 46, 49). Durepos et al (32) found that informal caregivers considered this ability to explore, access, and utilise the information and support services available to them as pivotal when maintaining a sense of control in their own lives. Informal caregivers firmly advocated for accessible and navigable guidance and resources to assist them during this turbulent period, preparing them for the decline and death of the PLwD they were caring for (23, 24, 28, 36, 42, 44, 59, 61).

#### Changing with the dynamic situation

Dementia trajectories have been labelled ‘unpredictable’, with informal caregivers citing their frustration and confusion when faced with changes to their own and their person living with dementia’s daily lifestyle, and consequent change in the dynamics of their relationship (35, 44). Informal caregivers highlighted appropriate support, resources, and digestible information provided by care professionals as fundamental to their understanding and subsequent ability to adapt to the changing landscape of dementia (32, 33, 42, 64, 66).

Informal caregivers in several studies narrated their experience of transitioning into a caregiving role, and adapting to the new parameters of their relationship with the person living with dementia (28, 36–38, 44, 53, 54). These participants noted the benefits of attending peer support groups, and observing the experiences of others going through a situation that resembled their own and seeing how other informal caregivers navigated their roles (29, 36, 38, 39, 63). These experiences enabled them to anticipate challenges in their own person’s care trajectory. Informal caregivers also noted the value of care-staff providing continual reassurance during periods of change and uncertainty (26, 27, 37, 44, 51, 52).

#### Observing care in practice

In thirteen articles, carers praised the value of observing dementia-care in practice to gain a deeper understanding of the condition, pertinent to their own caring role (23, 26, 28, 34, 40, 42, 43, 48, 49, 60, 64, 65, 67). In addition, observing care-staff demonstrating emotion during their tasks also promoted the emotional wellbeing of informal caregivers, and provided reassurance which alleviated the sense of burden and guilt associated with transitioning a person into residential care (34, 42, 43, 48, 64).

Studies found that where informal caregivers observed PLwD engaging in meaningful activities, their own understanding of dementia and sense of wellbeing improved. Lemos (41), Midtbust et al (47), and Slape (55) depict how such observations reassured informal caregivers in when deciding to transition their PLwD they were caring for into residential care. Participants came to a realisation that people can live well with dementia when witnessing such activities in practice, as opposed to merely being informed about them by care-staff (41, 47, 55).

### Theme 2: Engagement in care-based decision making

This theme embodies the sub-themes of “being valued by care professionals as an expert on the person they were caring for”, “reducing the sense of obligation”, “appropriately timed conversations”, and “making ‘involvement’ accessible”. These sub-themes reflect a narrative presented in the literature, establishing the preferences of informal caregivers when engaging or rejecting opportunities to be involved in care-based decision making. These sub-themes portray nuance within informal caregiver experiences, where lived experiences and dynamics had moulded their attitudes and beliefs towards the care sector.

#### Being valued by care professionals as an expert on the person they were caring for

Significantly, twenty-one studies identified that informal caregivers wished to be consulted when professional care teams were making care-based decisions for PLwD they were caring for, irrespective of the dynamic between the informal carers and those living with dementia (21, 23, 24, 26, 27, 30, 34, 36, 38, 40, 42, 44, 45, 48, 50, 56, 60, 62, 64–66). However, some studies highlighted informal caregivers’ preferences to disengage with the decision-making process for their own wellbeing, despite appreciating opportunities to play an advocacy role (35, 46, 54, 61). Harrad-Hyde et al (61) particularly showcased families’ wishes to disengage in this process, and a need for care-staff to respect this decision and not intrude during emotionally sensitive times.

Informal caregivers held an important advocacy role, but reported a spectrum of experiences in which the level of respect and empowerment from care-staff varied significantly. Informal caregivers demonstrated a keen awareness of what care professionals considered “best care” for PLwD, but iterated a need to be respected in their decisions to provide “comfort care” which might conflict with what professionals recommended, such as advocating for a person’s decision to not take symptomatic medications (33, 42, 60). Moreover, informal caregivers reported the significance of empowerment by care-staff, referring to practices where care-staff enabled carers to gain confidence in their own care-based decisions and assisted them in overcoming feelings of guilt (23, 24, 27, 37, 47, 60).

Despite studies highlighting the need for cultural-competency training as a practical implication for care-staff (21, 32, 39, 45, 48), only Durepos et al (32) identified informal caregivers’ need for healthcare professionals to respect their cultural values when making care-based decisions.

#### Reducing the sense of obligation

Informal caregivers with varying levels of rapport with their those they were caring for reported a need for care-staff to take on the burden of caregiving (23, 44, 46, 47, 54). Notably, the literature highlights that their sense of obligation is only reduced when care-staff are knowledgeable about dementia (28, 43, 65). Care-staff ability to approach PLwD with certainty and confidence was considered a key indicator of competence, by informal caregivers (23, 46, 54, 60).

Informal caregivers reported feeling reassured by care staff taking responsibility for the care of PLwD, and by them demonstrating initiative in adhering to previous documentation and preferences during a crisis or incident (40, 44, 54). Expanding on these points, Khemai et al (40) noted that appropriate use of documentation avoided the need for informal caregivers to repeat themselves and to interject when the person they were caring for was in crisis.

#### Appropriately timed conversations

Informal caregivers expressed a need for care-staff to be proactive in initiating conversations surrounding the care planning of PLwD (27, 39, 40, 45, 47, 60). Fifteen studies stressed a need by informal caregivers for these conversations to be timely, and in advance of a crisis or incident involving PLwD (21, 27, 30, 33, 35, 36, 40, 44, 45, 50, 51, 53, 56, 60, 63). Timely conversations were perceived to have multifaceted benefits, regardless of informal caregivers’ preferences to engage with the care-based decision-making process. Informal caregivers who spoke retrospectively, and who had not been involved in care-planning, lacked confidence when having to initiate care-based conversations: citing uncertainty about who was the appropriate point of contact (33, 40, 53, 60).

Informal caregivers, in research, spoke of the need to be encouraged by care-staff to consider the prognosis of PLwD and the impact that this might have on them in the future. Here, participants highlight the importance of being provided with repeated opportunities to revisit care-based decisions or become involved in care-planning processes, even if previously declined (27, 30, 39, 45, 64). Interestingly, no studies reported a counter-perspective, with all eligible studies voicing informal caregivers’ wishes to be provided repeated opportunities to engage with the care-planning process.

#### Making ‘involvement’ accessible

Participants stressed the significance of care-staff and the wider sector using lay-language when conveying information (29, 31, 36, 46, 49). Multiple studies reported themes reflecting a sense of hopelessness amongst informal caregivers and feeling overwhelmed, which were strongly attributed to the use of technical jargon by care professionals (34, 51).

Givens et al (34), Mclennon et al (46), and Wladowski (57) also described the need for care professionals to support and explain paperwork. They highlighted the frustration and confusion that informal caregivers felt when faced with an unending flow of paperwork, without explanation and support in clear and simple language. Before being asked to make care-based decisions, informal caregivers voiced a need to be provided with adequate information by care professionals. Retrospectively and in real-time, informal caregivers iterated the importance of understanding the potential benefits and consequences of care-based decisions, prior to being asked to engage in decision-making processes (21, 27, 29, 33, 34, 36, 40, 44, 45, 48, 51, 56, 57, 60, 61, 64, 65). Being provided with information tailored to the needs of PLwD and information on alternative options, even if not recommended as “best care” by the care service, was particularly sought (35, 42, 52, 54).

### Theme 3: Coping mechanisms and support for own wellbeing

This theme outlines informal caregivers’ coping mechanisms to ensure their own physical and psychological wellbeing, in preparation to face the deterioration of the condition of the person they were caring for. Sub-themes of “rapport building with care-staff”, “the role of friends and family”, “understanding self through others”, “reframing perspectives” and “self-care” highlight the significant role of maintaining social relationships, nurturing those networks, and the importance of reflection and self-awareness during this experience.

#### Rapport building with care-staff

Informal caregivers noted the value of establishing relationships with care-staff, where care-staff could earn their trust, subsequently encouraging them to engage in the process of care-based decision making (21, 24, 27, 29, 31, 32, 35, 38, 42, 45, 49, 51, 55, 58, 60, 61, 63, 65). Two studies particularly highlighted this rapport as pivotal to promoting casual conversations between care-staff and informal caregivers, thus encouraging the families to consider care-based decisions and future planning in a relaxed environment (27, 42).

Informal caregivers perceived positive relationships with care professionals as an avenue to have frank and honest discussions. Trust and transparency from care-staff was described as key to easing anxiety and guilt (37, 60). Particularly, Daneau et al (31) observed that trust between informal caregivers and care-staff, and demonstrated how this led to greater transparency from care-staff if there had been an incident with the person they were caring for. When discussing rapport, informal caregivers noted how the continuity of care-staff was paramount to ensuring this trust (24, 32, 63). The continuity of care-staff was foundational to establishing trust and honesty, with themes indicating their need to utilise care-staff as an avenue for emotional support and understanding (27, 49, 55, 56).

#### The role of peer support

Informal caregivers iterated the importance of seeking and nurturing supportive networks amongst family and friends, alongside a rapport with care-staff. Though the term ‘anticipatory grief’ was not explicitly used by the eligible literature, some studies identified similar experiences within informal caregivers’ accounts, signifying the importance of reminiscing memories of the people they were caring for with peers who could also contribute their experiences with said-person, and provide emotional support (38, 41, 59).

Conversely, some participants emphasised their fear and anxiety surrounding the strain that this may place on their personal relationships (28). Strain was reportedly rooted within familial conflict, where informal caregivers sought a consensus from their and PLwD’s wider family, to collectively make care-based decisions (29, 44, 50). Whilst noting that interpersonal strain and anxiety may manifests during familial conflict, informal caregivers reported a sense of camaraderie in sharing the burden of caregiving (27, 35, 46, 62).

Whilst the adverse experience of navigating the dementia trajectory strengthened the relationships between friends and family of some participants (35, 46, 60), other papers iterated a need to seek support beyond familiar circles; highlighting the value of meeting individuals who may be going though similar experiences and confiding in a non-judgemental setting (28, 36, 39, 63). In seeking support beyond their familiar network, informal caregivers delineated how fear and anxiety surrounding interpersonal conflict and strain were alleviated. Six studies cited the use of dementia carer support groups and online forums as paramount sources of support, where they could share experiences and seek emotional support from individuals distant from their situation, but who could empathise with their experience (28, 29, 36, 38, 39, 63). By volunteering in dementia-based organisations, informal caregivers further showcased the value of connecting with people who had similar lived experiences and could understand the nuance and complexities of their context (28, 37).

#### Reframing perspectives

When discussing their evolution from a peer to caregiver for PLwD, participants noted the importance of reframing their perspective to support their own wellbeing. Five studies shared how participants utilised practices of faith or spirituality for guidance, leading to a deeper understanding to their situations (35, 39, 45, 46, 54). Particularly, McCarthy et al (45) conveyed how religion became a key tool for acceptance: enabling informal caregivers to accept the idea that some circumstances were beyond their control, whilst maintaining trust in divine intervention. Interestingly, Han et al (35) identified this within participants who had no prior religious nor spiritual beliefs, but had resorted to providence to find comfort and reassurance during the unpredictability of their person’s decline.

Irrespective of religious and spiritual practices, informal caregivers identified the importance of acceptance throughout their experience, especially coming to terms with their reality (42, 52). Rather, informal caregivers shared their acceptance of their person’s dementia trajectory as a terminal condition, framing it as a key driving force for consideration and planning for a future without the person they were caring for (33, 55, 65).

#### Self-care

Informal caregivers widely acknowledged how they needed to care for themselves, to effectively care for others. Scheduling time for their own hobbies and interests was often perceived as important to informal caregivers, as any other responsibilities they held, ensuring they were able to maintain an identity beyond their role as a caregiver (32, 33, 35, 63). Conversely, Carter et al (28) showcased the antithesis of this, with one participant citing their dependence on alcohol as a self-care practice.

Informal caregivers also voiced the difficult, yet necessary, action of disengaging with their role and relationship with PLwD for self-preservation. Some participants shared their difficult decisions to avoid or postpone their visits to the care setting, citing it as a source of distress, particularly when PLwD did not recognise them (24). Similarly, informal caregivers noted their gradual detachment from the situation entirely by acknowledging the change in dynamics and withdrawing from their personal relationship with the person they were caring for; articulating this as a necessary step towards overcoming anticipatory grief, and preparing for the next step in their lives (37, 38, 63).

## LIMITATIONS

### Limitations within the literature

Though eight of the eligible articles did not discuss any limitations to their work, the remaining literature highlights limitations in their studies which may inform future research designs and conduct.

Key limitations included methodological weaknesses, with studies acknowledging the limitations of conducting research in a single setting (24, 25), conducting a cross-sectional design with a single point of data collection (39, 43, 51, 54, 60), and using a self-selected sample (28, 30). Review papers noted limitations surrounding their inclusion criteria, with Harper et al (36) including articles published after 2000 and Cresp et al (29) only including studies published between 2010 to 2020. Interestingly, Bosco et al (65) stated that potential evidence may have been excluded from their review, with limitations surrounding the lack of specificity in reporting of qualitative methodologies.

Restricted generalisability was significantly noted across the literature, with papers acknowledging the role of context and culture in how care sectors and informal caregivers engage with certain practices. Three studies conducted in the USA (23, 33, 34), one in Canada (32), one in Israel (21), and one in Sweden (54) all highlighted the limited international generalisability of their findings. Brazil et al’s (58) multi-national study also cites the role and context of the COVID-19 pandemic restrictions as a key limitation to their project, impeding their ability to implement their proposed intervention.

Reflecting on their design and conduct, some articles noted the practical challenges surrounding research within this domain. With surrounding literature attesting to the challenge of accessing service-users and retaining participants throughout a research project (68), Thompson et al (56) cited care-staff turnover as a key limitation to their study. In a dyadic exploration of both care-staff and informal caregiver perspectives, Thompson et al (56) outlined the obstacles faced as care-staff would leave employment during the course of the study, thus withdrawing their participation.

Interestingly, both Caron et al (27) and Harrad-Hyde et al (61) suggest their opportunity sampling may have created bias. Caron et al (27) specifically cite their recruitment of participants who all maintained strong positive relationships with PLwD, highlighting that their findings may be skewed due to the overrepresentation of positive experiences of informal caregivers. Cho et al (59) similarly share the potential bias in their study, attributed to the involvement of researchers who were also conducting the parent study of this paper.

### Limitations of this review

Though this review rigorously adheres to the JBI methodology for scoping reviews (18), there remains limitations surrounding the review process which must be acknowledged.

To minimise subjectivity in the data extraction process, both reviewers (CM and JD) conducted full paper screenings to check eligibility of all papers, compared their decisions, and discussed any discrepancies to come to an agreement. Despite both reviewers (CM and JD) testing their initial level of agreement by screening the first ten papers independently, and comparing their decisions, the addition of a third reviewer may have improved reliability by mediating the discrepancies between CM and JD. The addition of other reviewers may also have increased the rigour and credibility of the conclusions, placing the data extraction process and subjective interpretation of findings under further scrutiny.

## CONCLUSION

The literature surrounding experiences of informal caregivers of PLwD in residential care settings is accumulating, though very few studies have taken a longitudinal approach to understand how their experiences and needs evolve over time.

Recurring themes identified informal caregiver needs of having a thorough knowledge and understanding of dementia, set expectations, and guidance from practitioners regarding decision-making and advance care planning to minimise distress during a crisis. Other prominent needs included clear and timely communication, trust, and rapport with care-staff, alongside access to both formal and informal networks who can provide comfort and emotional support whilst navigating the process.

When discussing preferences for their involvement in care-based decisions, informal caregivers identified a need to be seen as a valued member of the care team and as an expert, ensuring that PLwD’s wishes were respected whilst also alleviating the strain of being sole decision-makers. Informal caregivers’ preferences also highlight a need for certainty and concise information from care-staff in lay-terms, with care-staff also taking initiative to sensitively initiate timely conversations surrounding advance care planning. Though the extent of preferred involvement varied amongst participants, informal caregivers voiced a desire to be approached with repeated opportunities to revisit decisions as PLwD’s condition progressed, in a manner respectful of their circumstances.

Informal caregivers’ coping mechanisms to ensure their own wellbeing varied from balancing self-care practices, maintaining boundaries to minimise burnout, to overcoming isolation by seeking comfort and meaningful relationships with peers and the wider dementia-care community. Moreover, some informal caregivers resorted to faith or spirituality to establish a sense of purpose and attribute a deeper meaning to their circumstances.

This review highlights several implications for practice. Practitioners must prioritise a proactive, compassionate approach towards informal caregivers of PLwD to support them both emotionally and practically: providing resources and services to guide them from admission to the stages of end-of-life care. Beyond emotional support, practice should also make efforts to inform informal caregivers on the reality of dementia, their person’s trajectory, and terminology, to set expectations and prepare them before being asked to engage in any decision-making. To minimise confusion and ensure a sense of trust and stability for informal caregivers, care settings should also provide each person living with dementia’s circle of support with a designated point of contact upon admission.

Future research should consider a longitudinal approach to understand how the role of an informal caregiver evolves, how their experiences unfold as their person’s condition progresses, particularly examining the differences between engaged and disengaged informal caregivers. Studies should also explore the experiences of informal caregivers from underrepresented communities, and examine the needs and preferences specific to different cultural and ethnic backgrounds. Though findings highlight informal caregivers’ wish to be sensitively approached repeatedly for advance care planning discussions, future studies should specifically seek to understand what this preferred process looks like for informal caregivers. Further avenues also include an exploration into the influence of trust on care-based decision engagement, how the relationship between informal caregivers and PLwD regulates caregivers’ decisions, and understand how informal caregivers would like to be supported by professional services and what this would entail.

### Funding

The author(s) disclosed receipt of the following financial support for the research, authorship, and/or publication of this article: This PhD project is supported by the University Research Studentship Award (URSA) from the University of Bath.

### Patient consent for publication

Not applicable.

### Data availability statement

All data relevant to the study are included in the article or uploaded as online supplemental information.

### Ethics approval

Not applicable.

