## Supplementary Table 1 for "The First-Hand Needs of Informal Caregivers of People Living with Dementia, in Residential Care Settings: A Scoping Review"

| Citation | Location | Research Questions/Aims | Sample (relationship to the PLwD) | Identified needs of informal caregivers | Coping mechanisms or support mentioned to improve own health and wellbeing | Preferences and/or suggestions for how informal caregivers could be involved in the decisions and care of PLwD | Limitations | Implications for practice | Avenues for future research |
|---|---|---|---|---|---|---|---|---|---|
| Armstrong, Alliance, Corsentino, Maixner, Paulson & Taylor (2020) | USA | Investigate barriers to quality end-of-life care in the context of dementia with Lewy bodies (DLB) | Family caregivers<br><br>Interviews (N=30)<br><br>Survey (N=657) | Knowledge and understanding of their loved one's diagnosis.<br><br>Getting a firm diagnosis.<br><br>Receiving guidance and signposting to find a facility suitable to their and their loved one's needs.<br><br>Witnessing and communicating with care staff who demonstrate competence in dementia care.<br><br>Consistent staff throughout the course of loved one's care.<br><br>Setting expectations of the disease trajectory. | Rapport building with familiar care staff.<br><br>Knowing that dementia is a terminal condition. | Value family as a key contributor in care-based decisions within the care team, but not to fully offload the responsibility to the family.<br><br>Support families and reassure them in their decisions, regardless if not the choice of clinical staff. | Limited to US-based healthcare, limiting international generalisability. | Raise awareness and educate families of their loved one's diagnosis in early stages to set expectations.<br><br>Improve physician knowledge on dementia subtypes, and train physicians to communicate with families. | Identify DLB stages.<br><br>Recognition of end-of-life pathway in DLB<br><br>Factors that drive quality end-of-life experiences, and best practices for tailoring palliative approaches for DLB. |
| Ashton (2013) | UK | Understand Advance Care Planning process and experiences from the perspectives of family caregivers as proxy decision makers for PLwD at the end-of-life. | Family caregivers (N=12) | Knowledge and understanding of loved one's diagnosis, the trajectory, and how the disease may change them.<br><br>Continuity of care staff who build rapport with families.<br><br>Make information and support services easily accessible and navigable.<br><br>Prior knowledge of long-term care facilities, and end-of-life care prior to there being a crisis.<br><br>Communication with their loved one. | Setting expectations from care staff in the early stages.<br><br>Avoiding or postponing care visits to save upset from not being recognised by loved one.<br><br>Trusting relationships with care staff. | Support families through the decision-making process and reassure them in the decisions made.<br><br>View family as experts and consult them when making care-based decisions. | Study conducted within one specialist unit. | Training to ensure adequate inter-team collaboration.<br><br>Educate and train families on dementia diagnoses, trajectory, and subsequent behaviour management techniques. | develop care pathways specific to meet the needs of people with advanced dementia<br><br>Explore attitudes to advanced care planning.<br>3<br>Longitudinal study to demonstrate the impact of support, education and skills training on the grief anxiety. |
| Ashton, Roe, Jack & McClelland (2016) | UK | Explore the experiences of advance care planning amongst family caregivers of PLwD. | N=12<br><br>Spouse (*n* = 4)<br>Child (*n* = 4)<br>Niece (*n* = 2)<br>Granddaughter (*n* = 1)<br>Sibling (*n* = 1) | Knowledge and understanding of what advanced care planning means, and entails.<br><br>Open and honest discussions with care staff.<br><br>Staff viewing their loved one as a person rather than someone living with dementia. | Perceiving themselves as responsible for their loved one.<br><br>Consider advanced care planning as an opportunity for discussion and advocacy, rather than something negative.<br><br>Considering their loved one as a whole person and maintaining their sense of self. | Informal discussions with professional care staff<br><br>Facilitate advanced care planning discussions early on, to make inevitable decisions sooner. | Sample limited to one dementia specialist care unit. | Consider 'historical identity' of PLwD and carry their prior preferences and lifestyles into advanced care planning.<br><br>Don't assume family are experts of the PLwD. | Attitudes towards advanced care planning |

| Citation | Location | Research Questions/Aims | Sample (relationship to the PLwD) | Identified needs of informal caregivers | Coping mechanisms or support mentioned to improve own health and wellbeing | Preferences and/or suggestions for how informal caregivers could be involved in the decisions and care of PLwD | Limitations | Implications for practice | Avenues for future research |
|---|---|---|---|---|---|---|---|---|---|
| Black et al. (2009) | USA | How do surrogate decision makers for PLwD develop an understanding of patient preferences about end-of-life care and patient wishes? | Surrogate decision makers (N =34) | Observing experiences of other PLwD and their families in similar predicaments<br><br>Having trusting relationship between themselves and PLwD for effective decision-making.<br><br>Having end-of-life care and preference discussions with PLwD in advance of admission. | Having encouragement from care staff, reassurance in their decision-making | Being encouraged to consider and discuss the future with their loved one and care staff through official conversations, even if they don’t feel emotionally prepared.<br><br>Having difficult conversations early on; pre-admission if possible.<br><br>Having the opportunity and feeling included in advanced care planning. | Perceived wishes and preferences of PLwD couldn’t be checked for accuracy as PLwD were too cognitively impaired to participate. | Care staff should promote and facilitate difficult conversations between informal caregivers and PLwD to understand needs and preferences. | None discussed |
| Boogaard, Werner, Zisberg & van der Steen (2017) | Israel | Assess trust of family caregivers of PLwD in professional care staff in nursing homes, and possible correlates | Family caregivers (N=215) | Trusting relationships with physicians<br><br>Strong and timely communication from staff. | Building trust and rapport with professional care staff. | Collaborating with members of the professional care team who are native to the same ethnicity and culture as themselves.<br><br>Providing family caregivers with adequate information before encouraging care-based decision to be made. | A focus on Israeli care homes may limit generalisability. | Continuous communication with the family regarding PLwD health status is key. | Assess informal caregiver trust in particular activities of professional care staff.<br><br>Qualitative study looking at how trust could be improved or undermined. |
| Bosco et al. (2024) | UK | Conduct a systematic review of literature exploring the experiences of family carers of PLwD accessing hospice settings for end-of-life care. | 4 articles | Developing knowledge on pain management and providing comfort care.<br><br>Witnessing PLwD being supported by competent care staff.<br><br>Having expectations set by care professionals, to understand the process and available support for themselves.<br><br>Trusting relationships with care staff. | Understanding dementia as a progressive condition and accepting the unpredictability of the condition.<br><br>Remaining aware of their own limits and boundaries, to reduce stress created by their responsibilities.<br><br>Releasing sorrows to a support service provided by the care service.<br><br>Matching expectations within the circle of informal care of PLwD. | Being used as a key resource of information of PLwD, so their person can receive personalised care.<br><br>Professional care staff delivering information in-advance of decision making, discharge, and other key processes. | Potential evidence had been excluded due to a lack of specificity in methodology of papers. | Active involvement of family carers in care-based decisions of care teams.<br><br>Increase awareness surrounding in-patient hospice and end-of-life care for PLwD. | Explore the experiences specific to hospice dementia care.<br><br>Qualitative studies must provide rigorous reporting of research methodology.<br><br>Explore the experiences of underserved communities (e.g. minority ethnic groups, religious communities).<br><br>Share supplementary data supporting qualitative studies, so findings can be replicated in future.<br><br>Specify context of participants where quotations are included. |

| Citation | Location | Research Questions/Aims | Sample (relationship to the PLwD) | Identified needs of informal caregivers | Coping mechanisms or support mentioned to improve own health and wellbeing | Preferences and/or suggestions for how informal caregivers could be involved in the decisions and care of PLwD | Limitations | Implications for practice | Avenues for future research |
|---|---|---|---|---|---|---|---|---|---|
| Brazil et al. (2024) | Canada, Czech Republic, Italy, Netherlands, Republic of Ireland, and UK. | Assess the impact of an Advance Care Planning intervention on family satisfaction, decision-making, and identify costs and supportive conditions for the intervention. | Family carers (n=101) | Trusting relationship with care staff.<br><br>Working alongside care staff who demonstrated empathy, knowledge and skills specific to understanding their PLwD. | Support services and care planning which also prioritises the family.<br><br>Good communication with care staff. | None discussed. | Incomplete and inconsistent data collection.<br><br>Influence of COVID-19 pandemic guidelines on how intervention was designed to be delivered, and therefore results may differ under typical circumstances. | Provide training to ensure care staff are confident, comfortable, and capable of engaging families in end-of-life care discussions.<br><br>Care-staff should prioritise strengthening relationships with families, to promote their involvement. | Reassess the impact of said-intervention under circumstances where COVID-19 restrictions aren't in place.<br><br>Explore country-specific efficacy and whether context affects the implementation of intervention.<br><br>Explore family-staff communication dynamics. |
| Caron, Griffith & Arcand (2005) | Canada | Examine the experience and preoccupations of family caregivers about end-of-life issues, and treatment decision-making processes in the context of advanced dementia | Family caregivers (N=24) | A personalised relationship with the care providers.<br><br>Good first impressions from care staff.<br><br>Trusting and supporting relationship with care staff.<br><br>Regular and timely communication from care staff, and expectations being set by the care providers to avoid disappointment.<br><br>Knowledge on the trajectory of their loved one's disease. | Emotional support from care staff.<br><br>Meeting regularly for updates from care staff and being informed in terms easily understood.<br><br>Reassurance and dispelled doubts by care staff. | Offering informal conversations as a comfortable platform to discuss serious issues and care decisions.<br><br>Repeatedly offering an opportunity for families to discuss and re-visit care decisions.<br><br>Providing information for families before pressing them to make decisions.<br><br>Empower families to feel competent in their ability to inform shared decisions on care. | Study focusses on families with good relationships with their loved ones. | Be proactive and approach families to provide opportunities for them to ask questions, get information, and re-visit care-based decisions.<br><br>Approach interventions or decision influencing with sensitivity to families' feelings. | See how the relationship between informal caregiver and PLwD influences decision making. |

| Citation | Location | Research Questions/Aims | Sample (relationship to the PLwD) | Identified needs of informal caregivers | Coping mechanisms or support mentioned to improve own health and wellbeing | Preferences and/or suggestions for how informal caregivers could be involved in the decisions and care of PLwD | Limitations | Implications for practice | Avenues for future research |
|---|---|---|---|---|---|---|---|---|---|
| Carter et al. (2018) | UK | Explore the experiences and preparedness of family caregivers on best interest decision-making of a loved one living with advanced dementia | N=20<br><br>Parent (n=13)<br>Sibling (n=4)<br>Spouse (n=2)<br>Extended family (n=1) | Seeing their loved one safe, and happy in the care setting<br><br>Strong social support circles being maintained by friends and family under the strain of the situation.<br><br>Care staff being knowledgeable on dementia care.<br><br>Care staff being familiar with themselves and their loved one's preferences.<br><br>Clear signposting for support services | Over-reliance on alcohol for wellbeing<br><br>Confiding in friends and family where possible, but fearful that it puts strain on relationships.<br><br>Volunteering within a dementia organisation to meet others going through similar experiences and feel rewarded. | Becoming a substitute decision-maker for their loved one feels like a natural progression in role and gives sense of self-worth. | Self-selected sample who were enrolled on an Advanced Care Planning intervention | Enhance care staff expertise on dementia care practices, for better care delivery and reassurance for informal caregivers. | Develop a family carer psycho-educational care intervention |
| Cho, Sefcik, Washington, Oliver & Demiris (2024) | USA | Understand caregivers' social support whilst PLwD are being enrolled into hospice care. | N=22<br><br>Child (n=10)<br><br>Spouse (n= 4)<br><br>Significant other (n=6)<br><br>Relatives (n=2) | Demonstrations of support from professional care staff and peers.<br><br>Accessible and easily navigable support services.<br><br>Consistency of support services and communication from hospice. | Relying on peer support from family and friends, specifically practical assistance which enabled them to take breaks from their caregiving responsibilities.<br><br>Maintaining connection with the PLwD.<br><br>Sharing their feelings and experiences with trusted peers who can empathise. | None discussed. | Potential bias from researchers who were involved in the parent study. | Consider the social support of caregivers during the enrolment process.<br><br>Assess the needs of family caregivers whilst assessing PLwD for care enrolment. | Explore factors influencing caregivers' access to support.<br><br>Understand the characteristics of supportive relationships between families and professional care staff. |

| Citation | Location | Research Questions/Aims | Sample (relationship to the PLwD) | Identified needs of informal caregivers | Coping mechanisms or support mentioned to improve own health and wellbeing | Preferences and/or suggestions for how informal caregivers could be involved in the decisions and care of PLwD | Limitations | Implications for practice | Avenues for future research |
|---|---|---|---|---|---|---|---|---|---|
| Cresp, Lee & Moss (2020) | Australia | Conduct a systematic review to see how substitute decision-makers are affected by and experience making decisions for PLwD at end-of-life. | 7 articles | Trusting relationships with care staff.<br><br>Staff setting expectations early on regarding the constraints of what they 'can' and 'can't' do when caring for PLwD<br><br>Certainty and understanding of what end-of-life looks like in PLwD. | Rapport with professional care staff provides emotional and physical support.<br><br>Diffusing familial conflict when having to collectively make decisions on their loved one's care.<br><br>Joining dementia support groups for advice and inform decision-making. | Care home staff must earn the trust of informal caregivers to encourage their involvement in decision-making.<br><br>Fully inform informal caregivers of the impact/outcomes of decisions before said decisions are made. | Limited to studies published between 2010-2020. | Staff must build trust with informal caregivers to provide them confidence in their care of PLwD.<br><br>Signpost informal caregivers to clear resources which educate on end-of-life care, and what they should expect.<br><br>Staff should assist informal caregivers in decision making by translating information into lay terms. | Explore how informal caregivers are affected by making decisions.<br><br>Examine how informal caregivers experience trust and mistrust.<br><br>Develop interventions aimed at reducing guilt, mistrust and confusion |
| Crowther (2011). | UK | Explore needs of PLwD and their informal caregivers in the last year of life and surrounding death | Informal caregivers (N=41) | Thorough and timely communication from professional care staff.<br><br>Knowing that the trajectory makes PLwD needs and preferences dynamic and changing | None discussed. | Informal caregivers to be considered as experts in the care of PLwD.<br><br>Provide informal caregivers a platform and repeated opportunities to engage in discussion.<br><br>Making decisions based on the personhood of PLwD, and their self before dementia. | Retrospective nature.<br><br>Self-selected sample. | Observe pre- and post-death grief and support families during this transition.<br><br>Educate care staff on the end-of-life experiences of PLwD. | Study the views and experiences of families across different cultures and ethnicities.<br><br>Explore experiences of PLwD of different sexualities, and queer communities who have 'chosen' families.<br><br>Explore and identify ways informal caregivers would like to be supported. |

| Citation | Location | Research Questions/Aims | Sample (relationship to the PLwD) | Identified needs of informal caregivers | Coping mechanisms or support mentioned to improve own health and wellbeing | Preferences and/or suggestions for how informal caregivers could be involved in the decisions and care of PLwD | Limitations | Implications for practice | Avenues for future research |
|---|---|---|---|---|---|---|---|---|---|
| Daneau, Bourbonnais & Legault (2022) | Canada | Propose a theory on nurses' supporting relatives who must make end-of-life care decisions of PLwD in long-term care homes. | N= 19<br><br>Nurses (n=9)<br>Relatives (n=10) | Strong and trusting relationship between informal caregivers and the professional care staff caring for PLwD.<br><br>Transparency from care staff on incidents or accidents | Regular conversations with care staff to clarify any uncertainty on their loved one's state. | Appropriately timed conversations to discuss care planning and set expectations.<br><br>Reduce need for informal caregiver intervention when there is an incident.<br><br>Care staff being proactive and taking initiative, informing themselves by previously set care plans and agreements.<br><br>Using plain language to deliver information. | No participants had poor relationships with care staff. | Nurses must work to develop trusting relationships with informal caregivers.<br><br>Reduce fears and anxiety of informal caregivers by providing lay term education on dementia in end-of-life stages. | None discussed. |
| Durepos, Ploeg, Sussman, Akhtar-Danesh & Kaasalainen (2020) | Canada | Explore the end-of-life experiences of caregivers of PLwD to understand factors perceived as influencing preparedness for death. | Informal caregivers (N=16) | Witnessing deterioration of loved one prepares family for death.<br><br>Knowledge and understanding of dementia, and the changes that will happen.<br><br>Transparent, collaborative and trusting relationships with care staff.<br><br>Continuity of staff. | Educating self to maintain a sense of control in the situation.<br><br>Planning, organising and adapting plans in line with the fluid situation.<br><br>Emotional support from care staff, viewing rapport as friendship and personal.<br><br>Reframing and making the best of the situation.<br><br>Practicing self-care and learning to regulate own emotions. | Respect cultural and moral standpoints of families when they're encouraged to engage in decision-making. | Data collected retrospectively.<br><br>Sample not representative of wider population. | Provide practical and emotional support to families.<br><br>Train care staff in active listening and diffusing emotional distress when managing families.<br><br>Staff should be transparent with information sharing. | Develop and advocate the impact of interventions on caregiver death preparedness. |

| Citation | Location | Research Questions/Aims | Sample (relationship to the PLwD) | Identified needs of informal caregivers | Coping mechanisms or support mentioned to improve own health and wellbeing | Preferences and/or suggestions for how informal caregivers could be involved in the decisions and care of PLwD | Limitations | Implications for practice | Avenues for future research |
|---|---|---|---|---|---|---|---|---|---|
| Forbes, Bern-Klug & Gessert (2000) | USA | Describe families' decision-making processes regarding end-of-life treatments for nursing home residents with moderately severe to very severe dementia | N=28<br><br>Daughters (n=10)<br>Wives (n=4)<br>Husbands (n=4)<br>Daughter-in-law (n=3)<br>Sons (n=2)<br>Sisters (n=2)<br>Nephew (n=1)<br>Sister-in-law (n=1)<br>Grandson (n=1) | Separating time from caregiving role to do things for themselves.<br><br>Accepting death as the outcome.<br><br>Staff to set expectations and prepare them for the trajectory of dementia.<br><br>Consistent and timely communication with professional care staff.<br><br>Knowledge and awareness of terminology surrounding end-of-life care and terminal illness. | Professional care staff reducing the sense of obligation for families to visit daily.<br><br>Schedule time for their own interest.<br><br>Self-care through sleep and eating habits.<br><br>Acknowledging you must look after yourself before being an effective informal caregiver. | Provide supportive guidance to encourage future planning.<br><br>Begin end-of-life care decisions with small, simple decisions before getting to more sensitive aspects (food preferences, to DNR).<br><br>Respect informal caregivers desire for comfort care-based decisions.<br><br>Provide necessary information before having to make decisions. | Limited sample according to location, limiting generalisability. | Have a consistent point-of-contact for family members to communicate with, and build trusting rapport.<br><br>Staff training to provide a comfortable safe space to address topic of inevitable death and dying of PLwD. | Interventions into advanced care planning<br>. |
| Givens, Lopez, Mazor & Mitchell (2012) | USA | Understand the sources of stress for family members of PLwD with advanced dementia in nursing homes | Family members (N=16) | Witnessing staff emotional involvement when delivering care.<br><br>Seeing staff respect patients of all stages of dementia.<br><br>Strong communication from physicians. | Emotional support from nursing home staff during the admission process. | Give informal caregivers sufficient information on the trajectory of dementia, before they decide advanced care plans.<br><br>Not to 'overwhelm informal caregivers with paperwork, without explaining everything for them to understand.<br><br>Consider the family members as equals to care team in the decision-making processes. | Study depended on subject recall, after some of their loved ones with advanced dementia had already passed away.<br><br>Boston based, with majority white female participants which may limit generalisability. | Provide families with sufficient information before encouraging them to make decisions on care. | None discussed. |

| Citation | Location | Research Questions/Aims | Sample (relationship to the PLwD) | Identified needs of informal caregivers | Coping mechanisms or support mentioned to improve own health and wellbeing | Preferences and/or suggestions for how informal caregivers could be involved in the decisions and care of PLwD | Limitations | Implications for practice | Avenues for future research |
|---|---|---|---|---|---|---|---|---|---|
| Gonella et al. (2023) | Italy | Explore family caregivers' of PLwD's experiences of communication with nursing home staff during the COVID-19 pandemic from admission to end-of-life. | N = 25<br><br>Niece/Nephew, Adult Child, Son/Daughter-in-law | Frequent, simple and transparent communication from care staff.<br><br>Regular and timely updates from care staff regarding their PLwD.<br><br>Care settings to shift towards digital approaches for communication with families who are long-distance. | Sharing decision-making processes with peers and professional care staff to overcome feelings of guilt and betrayal.<br><br>Witnessing professionalism and competence from care staff.<br><br>Remaining informed of dementia, specifically regarding their person's deterioration. | Approachable and friendly care staff encourages participation from family members.<br><br>Allow time for family members to make care-based decisions.<br><br>Provide families with adequate information before asking them to make decisions.<br><br>Respecting families' decisions for comfort-care, over recommendations of care staff.<br><br>Allowing families to have access to information, and access to the care setting to promote trust. | Cross-sectional design couldn't capture the dynamics of the experiences, individuals involved, and changes.<br><br>Length of PLwD stay was not reported. | Identifying a single point-of-contact for families to liaise with regarding their person's health and deterioration.<br><br>Promote discussions and encourage families to think about care-based decisions at the point of admission.<br><br>Consider digital forms of communication for frequent contact with families who are not physically present. | Understand the information needs of family caregivers.<br><br>Explore efficacy of different communication methods between care settings and family caregivers to reduce distress of families. |
| Han, Chi, Han, Oliver, Washington & Demiris (2019) | USA | Identify challenges, possible solutions that are resources for resilience, and expected consequences faced by family members of PLwD in hospices. | Family members (N=39) | Knowledge on how to manage own anxiety and emotional distress.<br><br>Creating tactile approaches to communicate and maintain a relationship with PLwD.<br><br>Choosing a care facility which feels homely for themselves and loved one.<br><br>Effective communication from care staff, with regular and timely follow-ups. | Accepting the unpredictable nature of dementia.<br><br>Reinforcing belief in themselves, and self-appraisal.<br><br>Maintaining a healthy lifestyle.<br><br>Trusting relationships with care staff.<br><br>Meeting with family to share decision-making burden and ideas to engage with their loved one.<br><br>Resorting to a faith or spirituality. | Relive pressure from family to be the sole decision-makers and encourage collaboration between families and the professional care team.<br><br>Provide specific information on PLwD diagnosis and prognosis before decisions need to be made. | Data extracted from a parent larger study. | Build trusting relationships with families. | Design interventions to facilitate access to supportive resources for caregivers' resilience.<br><br>Explore challenges to improving the quality of care by health-care providers.<br><br>Test the efficacy and effectiveness of supportive behavioural and coping interventions, designed specifically for caregivers. |

| Citation | Location | Research Questions/Aims | Sample (relationship to the PLwD) | Identified needs of informal caregivers | Coping mechanisms or support mentioned to improve own health and wellbeing | Preferences and/or suggestions for how informal caregivers could be involved in the decisions and care of PLwD | Limitations | Implications for practice | Avenues for future research |
|---|---|---|---|---|---|---|---|---|---|
| Harper et al. (2021) | N/A | Conduct a scoping review of the experiences, priorities, and perceptions of informal caregivers of nursing home residents with dementia | 114 articles | Good, regular and timely communication from staff<br><br>Agreement from all family members/loved ones involved in the decisions of PLwD care. | Prioritising physical communication with their loved one (e.g., hand holding) to maintain relationship.<br><br>Rapport building with the care staff.<br><br>Involvement in support groups, as opposed to family and friends.<br><br>Redefining their role as caregiver, to give them purpose after transitioning to a facility | Care staff should acknowledge their expertise in the care of their loved one.<br><br>Use lay terms when navigating through complex information and procedures, making the knowledge accessible.<br><br>Personal-centred care being informed by themselves. | Only included studies published in English.<br><br>Limited search to peer reviewed literature published after 2000. | Consider informal caregivers as part of the care team.<br><br>Use lay terms when communicating and informing informal caregivers.<br><br>Building trust with informal caregivers will encourage their willingness to seek care for PLwD | Develop a tool to assess caregiver satisfaction.<br><br>Stakeholder pragmatic trials to understand barriers and facilitators of delivering interventions. |
| Harrad-Hyde, Jones, Agarwal, Faull & Birt (2024) | UK | Explore the potential role of peer-mentors when supporting families to prepare for care-based decision discussions. | N =29<br><br>Current family member (n=14)<br><br>Bereaved family member (n=15) | Practical support to navigate the system.<br><br>Vent emotions to a confidant to avoid feeling overwhelmed.<br><br>Acknowledge and understand that the person will deteriorate.<br><br>Developing a trusting relationship with care-staff early into their care journey.<br><br>Information to make informed decisions in PLwD best interests.<br><br>A balance of formal and informality with relationships between themselves and care staff. | One-to-one peer support from friends and colleagues who could relate through similar situations.<br><br>For peers and professionals to not intrude during the sensitive time, and respect the family's privacy. | Provide a platform for families to normalise and have these discussions about care and deterioration.<br><br>Remain respectful of families' decision to not engage during this sensitive time.<br><br>Provide and signpost families to necessary information before asking them to make decisions. | Opportunity sampling may cause selection bias. | Consider creating a peer-mentor role to liaise with families and build trusting relationships. | Explore avenues for how a peer-mentor program could be implemented in this context.<br><br>Develop and evaluate a peer-mentor program in care settings. |

| Citation | Location | Research Questions/Aims | Sample (relationship to the PLwD) | Identified needs of informal caregivers | Coping mechanisms or support mentioned to improve own health and wellbeing | Preferences and/or suggestions for how informal caregivers could be involved in the decisions and care of PLwD | Limitations | Implications for practice | Avenues for future research |
|---|---|---|---|---|---|---|---|---|---|
| Hennings, Frogartt & Payne (2013) | UK | Explore the caregiving experiences of spouse carers of people with advanced dementia living in nursing homes | Spousal Caregivers (N=10) | Having clarity on their new role, and what is expected or required of them.<br><br>Someone to confide in beyond their friend groups to avoid 'offloading' onto them and distancing friendships. | Attempting to detach themselves from the situation, to try and care less.<br><br>Compromising number of visits to balance their needs and wellbeing.<br><br>Volunteering with dementia organisations to build a friendly support network. | Reassurance and comfort to overcome feelings of guilt when making decisions. | None discussed | None discussed | Study conducted over a longer period of time. |
| Hennings & Froggatt (2019) | North America, UK, Australia | Conduct a narrative review to identify what is known about family caregivers' experiences of having a relative living with advance dementia in a nursing home. | 12 articles | Being recognised by their loved one.<br><br>Building relationships with nursing staff<br><br>Establishing new roles for themselves | Giving time to maintain the relationships with other family members and friends, to avoid loneliness.<br><br>Joining in carer support groups.<br><br>Support from nursing staff for informal caregivers during the transition of their roles<br><br>Disengaging with the process of losing the person. | Let caregivers play an advocacy role. | None discussed | Move away from 'carer' term and refer to the informal caregiver with a specific reference to their relationship with the PLwD.<br>Involve caregivers in decisions regarding PLwD care, treating them like a valued asset to the team. | Longitudinal study to explore the dynamic process.<br><br>Explore and compare experiences of engaged caregivers and disengaged caregivers who have severed ties with PLwD. |

| Citation | Location | Research Questions/Aims | Sample (relationship to the PLwD) | Identified needs of informal caregivers | Coping mechanisms or support mentioned to improve own health and wellbeing | Preferences and/or suggestions for how informal caregivers could be involved in the decisions and care of PLwD | Limitations | Implications for practice | Avenues for future research |
|---|---|---|---|---|---|---|---|---|---|
| Hill, Mason, Poole, Vale & Robinson (2017) | UK | Identify the end-of-life care priorities for informal caregivers and PLwD | N= 57<br><br>PLwD (n=14)<br>Informal caregivers (n=21)<br>Bereaved caregivers (n=22) | Maintaining close relationship with PLwD. | None discussed. | Planning care decisions ahead of time.<br><br>Be given the option to be involved in care decisions.<br><br>Not having care plans, and questions "pushed in [their] face". | Small sample size.<br><br>Data saturation was not reached | Consider a person-centred approach to family support.<br><br>Shared decision making between informal caregivers and professionals. | Repeat study with larger cohorts for full data saturation. |
| Hovland & Kramer (2019) | USA | Explore how caregivers handle dementia deaths, including identification of barriers and facilitators to preparing caregivers for the death of elderly PLwD. | Family caregivers (N=36) | Direct and regular communication from staff regarding prognosis and health status.<br><br>Direct and frank discussions with staff to set expectations.<br><br>Knowledge and understanding of the disease trajectory.<br><br>Having a certain prognosis.<br><br>Witnessing the decline of loved one prepares them for death. | Staff being proactive in informing families, to save them chasing everything.<br><br>Joining a support group of other families who have been through similar experiences.<br><br>Finding comfort through faith or engaging in rituals.<br><br>Believing in an afterlife to prepare them for death of their loved one.<br><br>Reading and educating themselves on dementia, and end-of-life care. | Offer repeated opportunities for family to engage in care-based decision discussions, for when they are ready to accept the situation.<br><br>Approach conversations with families sensitively. | Retrospective study where views may have been influenced over time after death.<br><br>Only one data collection point. | Staff need to be proactive in offering information regarding diagnosis, trajectory and end-of-life process.<br><br>Do not make assumptions on best ways to comfort families during end-of-life stages.<br><br>Cultural competency training for care staff. | Investigate how the experience of preparedness might unfold over time.<br><br>Understanding the experience of preparedness among multiple family members of persons with dementia.<br><br>Explore experience of African American caregivers' preparedness for death |

| Citation | Location | Research Questions/Aims | Sample (relationship to the PLwD) | Identified needs of informal caregivers | Coping mechanisms or support mentioned to improve own health and wellbeing | Preferences and/or suggestions for how informal caregivers could be involved in the decisions and care of PLwD | Limitations | Implications for practice | Avenues for future research |
|---|---|---|---|---|---|---|---|---|---|
| Khemai et al. (2022) | Netherlands | Examine the experiences of informal caregivers of PLwD with interprofessional collaboration among healthcare professionals. | N=32<br><br>Spouses (n=4)<br>Children (n=23)<br>Siblings (n=1)<br>Cousins (n=3)<br>Friends (n=1) | Regular updates from care staff regarding the health of their loved one.<br><br>Information on dementia, end-of-life and palliative care, and general information on nursing home life.<br><br>Timely communication from care staff.<br><br>Having a first point of contact | Monitoring and observing the care received by their loved one.<br><br>Guidance and support from professional staff, emotionally/physically/psychologically through the process of care home transition.<br><br>Witnessing communication and collaboration between healthcare team members. | Provide informal caregivers with all information before asking them to make care decisions.<br><br>Listen to informal caregivers when they relay personal information and preferences of PLwD, to tailor their care.<br><br>Document information and decisions made by informal caregivers, so they don't have to keep repeating themselves.<br><br>View and value informal caregivers as experts.<br><br>Be proactive and initiate decision-based conversations with informal caregivers, as they may be apprehensive. | Selection bias from nurses who recruited participants. | Provide informal caregivers with the necessary support, enabling them to feel comfortable in active decision-making.<br><br>Have a first point-of-contact for informal caregivers to establish a trusting relationship with and be provided with the necessary and timely information.<br><br>Be proactive to initiate timely decision-making conversations. | Compare experiences of all stakeholders within this experience of decision making. |
| Lemos (2023) | Netherlands | Explore the co-existence of experiences of anticipatory grief and manifestations of care to maintain meaningful relationships between informal caregivers and PLwD. | Ethnographic fieldwork-participant information not discussed | Regular discussions with care staff to keep up with dynamic changing needs.<br><br>Appreciating the contact and communication with their loved one with dementia.<br><br>Using meaningful objects to engage with PLwD (e.g., wine). | Finding meaning in relationships.<br><br>Maintaining meaningful relationships.<br><br>Making the best of the situation and finding alternative ways to connect with PLwD through adapting old hobbies. | Discuss loss of capacities between informal caregiver and physicians to firmly agree and establish trajectory of condition.<br><br>Collaborator by experimenting with art and relationships, with a focus to maintain connections and personhood of PLwD. | None discussed | None discussed | None discussed |

| Citation | Location | Research Questions/Aims | Sample (relationship to the PLwD) | Identified needs of informal caregivers | Coping mechanisms or support mentioned to improve own health and wellbeing | Preferences and/or suggestions for how informal caregivers could be involved in the decisions and care of PLwD | Limitations | Implications for practice | Avenues for future research |
|---|---|---|---|---|---|---|---|---|---|
| Lewis (2014) | USA | What is the lived experience of caregivers seeking hospice care or other formal end-of-life care for loved ones with dementia at the end of | Primary caregivers (N=11) | Guidance when choosing a care facility.<br><br>Rapport and trust with care staff to improve the care received by loved ones.<br><br>Communication and updates given in ways for everyone to understand.<br><br>Family education on dementia, end-of-life care, and what to expect.<br><br>Witnessing their loved one being cared for, 'beyond just being safe'.<br><br>Care staff to set realistic expectations and avoid optimistic statements. Signposting to navigate the process.<br><br>Physicians being actively involved in care and communication. | Transitioning from a mindset of seeking best care, towards seeking ways to make their loved one most comfortable.<br><br>Informal conversations with carers to prepare with the coming loss.<br><br>Good hospice carers take the pressure off having to visit daily, so time can be spent to care for themselves. | Allow family to make decisions for their loved one's comfort, rather than pushing for unwanted clinical intervention.<br><br>Promoting opportunities early on to make advanced care plans, to relieve guilt and stress later.<br><br>Make family feel heard and include them in conversations- valuing their expertise and viewpoint as advocates for PLwD.<br><br>Provide families with information on care alternatives before seeking decisions from them. | None discussed. | Improve support for caregivers in providing guidance and signposting throughout, from pre-admission to post-death grief.<br><br>Avoid comforting touch and empathetic statements which could trigger emotional release, only acceptable when the informal caregiver is already emotional and distressed. | Quantitatively determine the prevalence of these issues in caregivers of PLwD.<br><br>Explore pathways and experiences to seeking help in end-of-life stages. |
| Lopez, Mazor, Mitchell & Givens (2013) | USA | Explore family perspectives on person- and family-centred EoL care of nursing home residents with dementia | Family members of PLwD (N=16) | Witness PLwD receiving good quality care.<br><br>Sense of belonging between PLwD and staff, and informal caregivers and staff | Reassurance from professional staff regarding PLwD safety. | Clear, prompt and regular updates on loved one's condition, to make informed choices if not local | Does not reflect changes in perceptions over time. | None discussed | Exploring the hierarchy of needs from PLwD and informal caregiver perspectives<br><br>Explore intervention to enhance sense of safety to reduce stress and anxiety |

| Citation | Location | Research Questions/Aims | Sample (relationship to the PLwD) | Identified needs of informal caregivers | Coping mechanisms or support mentioned to improve own health and wellbeing | Preferences and/or suggestions for how informal caregivers could be involved in the decisions and care of PLwD | Limitations | Implications for practice | Avenues for future research |
|---|---|---|---|---|---|---|---|---|---|
| Lord, Livingston & Cooper (2015) | N/A | Conduct a systematic review explore barriers and facilitators to family carers of PLwD making proxy decisions, and interventions used to facilitate decision-making. | 30 articles | Signposting to available and alternative forms of support.<br><br>Timely information from staff regarding health status, and trajectory of loved one's condition.<br><br>Being able to involve PLwD in planning whilst they still have capacity.<br><br>Receiving reassurance about decisions they have made.<br><br>Knowing their loved one's previous wishes and experiences. | Acknowledge and adapt to changing role as caregiver.<br><br>Gaining an agreed family consensus before making decisions.<br><br>Overcoming family conflict.<br><br>Making care-based decisions before being faced with a crisis.<br><br>Influence decisions by considering what will reduce burden on the family.<br><br>Listening to the opinions of others, professional and personal relations. | Make families feel valued and listened to when raising concerns and when making efforts to engage them in decision making.<br><br>Promote decision making with family being perceived as members of the care team.<br><br>Encourage and provide multiple opportunities for families to make decisions prior to crisis.<br><br>Provide necessary information relevant before making decisions. | Most studies used convenience or purposive samples. | None discussed. | Develop and evaluate decision-aids for families. |
| McCarthy et al. (2023) | USA | Explore the perceptions and experiences of Black and White proxies of PLwD. | Proxies (N=44) | Knowledge and understanding of their loved one's diagnosis, what 'end-of-life' care and other terminology truly means.<br><br>Understanding what end-of-life stages look like in dementia.<br><br>Discussing needs and preferences with their loved one whilst they can.<br><br>Communication and timely updates on the condition of their loved one, in language understood. | Prioritising quality-of-life over clinical care for loved ones, even if it goes against recommendations from staff.<br><br>Trusting relationships with the care staff.<br><br>Care staff demonstrating knowledge of the PLwD.<br><br>Using religion as guidance and trusting that the situation is out of their own control. | Provide proxies with information prior to letting them make care-based decisions and advanced care planning.<br><br>Encourage advanced care planning on initial meeting and initiate those conversations with families.<br><br>Supporting families during decision-making.<br><br>Providing repeated and regular opportunities to discuss advanced care planning, and change care plans according to the dynamic situation. | Disruption to typical care practices during COVID-19 may influence the experiences. | Offer spiritual support to families, but not making assumptions based on ethnicity or race.<br><br>Cultural-competency training to staff. | None discussed. |

| Citation | Location | Research Questions/Aims | Sample (relationship to the PLwD) | Identified needs of informal caregivers | Coping mechanisms or support mentioned to improve own health and wellbeing | Preferences and/or suggestions for how informal caregivers could be involved in the decisions and care of PLwD | Limitations | Implications for practice | Avenues for future research |
|---|---|---|---|---|---|---|---|---|---|
| McLennon et al (2021) | UK AND USA | Identify needs, concerns, and advice from the blogs of caregivers caring for PLwD at the end-of-life. | N=6<br>Women caring for their mothers (n=5)<br>Women caring for their husband (n=1) | Family presence.<br><br>Support from healthcare professionals during care transitions.<br><br>Clear and plain language when being told information.<br><br>Confidence and certainty from care staff. | Sense of teamwork and camaraderie between relatives.<br><br>Spirituality and religious beliefs to give meaning and new perspectives. | Less responsibility during care transitions to reduce burden of communicating with various healthcare personnel, and paperwork.<br><br>Respect pre-decided life or death decisions. | Individual experiences limit transferability.<br><br>Cannot verify whether data is from actual informal caregivers. | Implementation of dementia nurse navigators.<br><br>Inform and set expectations of the upcoming experience for informal caregivers. | Explore psychosocial educational interventions |
| Midtbust, Alnes, Gjengedal & Lykkeslet (2021) | Norway | Explore family caregivers' experiences with palliative care for a close family member with severe dementia in long-term care facilities. | Family caregivers (N=10) | Seeing their loved one engage in meaningful activities in the care setting. | Being reassured by healthcare professionals in decisions of reducing visitation for their own wellbeing.<br><br>Regular contact from healthcare professionals if there were updates on their loved one. | Be approached sensitively when asked to contribute in the decision making process.<br><br>Arrange and hold meetings to make decisions well in advance of the terminal stages of their loved one. | Caution of this study being another burden for informal caregivers in this difficult period of their lives. | Build trusting relationships with informal caregivers to enable them to transfer the sense of responsibility comfortably. | None discussed |
| Moore et al. (2017) | UK | Understand the experiences of carers of PLwD, exploring the links between mental health and experiences of end-of-life care. | Family caregivers (N=35) | Receiving regular updates regarding their loved one's health status.<br><br>Being informed early-on regarding the likely trajectory of dementia.<br><br>Having their decisions respected<br><br>Eliciting the preferences of PLwD when they are still able to voice those preferences. | Moving their loved one to a different setting when they themselves weren't happy with the care being delivered.<br><br>Witnessing and experiencing genuine care and compassion from care staff.<br><br>Being able to observe and monitor the care received by PLwD in the setting. | Reinforce timely and sensitive information in written format for informal caregivers to revisit and learn.<br><br>Provide sufficient information to adequately prepare informal caregivers on the process and to make informed decisions.<br><br>Revisit care-based decisions after a crisis.<br><br>Giving informal caregivers the opportunity to influence care decisions for their loved ones. | Potential researcher bias from rapport building.<br><br>Small sample size. | Acknowledge the grief of informal caregivers, and provide support to mediate the process of admission and post-death. | Explore any modifiable factors that can aid informal caregiver grief processes.<br><br>Explore how to better support carers to feel empowered and supported in decision making. |

| Citation | Location | Research Questions/Aims | Sample (relationship to the PLwD) | Identified needs of informal caregivers | Coping mechanisms or support mentioned to improve own health and wellbeing | Preferences and/or suggestions for how informal caregivers could be involved in the decisions and care of PLwD | Limitations | Implications for practice | Avenues for future research |
|---|---|---|---|---|---|---|---|---|---|
| Moore, Crawley, Fisher, Cooper, Vickerstaff & Sampson (2023) | UK | Identify strategies that help family caregivers manage pre-death grief. | Family caregivers (N=150) | Develop new skills to be a better carer.<br><br>Preserve own identity throughout the process, by maintaining own relationships and own hobbies.<br><br>Renegotiating the parameters of the relationship with the loved one.<br><br>Staff continuity to build trust.<br><br>Knowledge and understanding of dementia and its trajectories | Being able to reciprocate the care they received from their loved one living with dementia in the past.<br><br>Spending time with their loved one.<br><br>Meeting others who share similar experiences through support groups or research events.<br><br>Emotionally detach from their loved one living with dementia and grieve.<br><br>Accessing formal support through counselling, for a neutral and non-judgemental output.<br><br>Embrace their identity as an informal caregiver. | Staff continuity in who engages with and builds rapport with the family. | No cultural diversity in the sample.<br><br>Self-report measures only. | Addressing pre-death grief in interventions for informal caregiver wellbeing. | Develop new bereavement models to include pre-death grief. |
| Raymond, Warner, Davies, Iliffe, Manthorpe & Ahmedzhai (2014) | N/A | Conduct a literature review to understand the experiences of being a carer of PLwD at end-of-lie | 12 articles | Making difficult decisions early on regarding end-of-life care practices.<br><br>Witnessing respect and dignity of their loved one being maintained.<br><br>Strong communication from the professional care team. | Having difficult conversations with their loved one living with dementia, to understand and advocate for their preferences.<br><br>Observing compassionate care of their loved one during visits. | Planning for end-of-life with conversations well in advance.<br><br>Make time to consult family and provide them with sufficient information.<br><br>Value the expertise of informal caregivers when consulting them in care planning.<br><br>Provide information on the implications of treatment decisions before asking family/advocates to make care-based decisions. | None discussed. | Provide care staff with cultural competency training to effectively have difficult conversations with families of different backgrounds.<br><br>Staff should be aware of elder abuse. | Direct experiences of informal caregivers going through end-of-life stages with loved ones living with dementia. |

| Citation | Location | Research Questions/Aims | Sample (relationship to the PLwD) | Identified needs of informal caregivers | Coping mechanisms or support mentioned to improve own health and wellbeing | Preferences and/or suggestions for how informal caregivers could be involved in the decisions and care of PLwD | Limitations | Implications for practice | Avenues for future research |
|---|---|---|---|---|---|---|---|---|---|
| Rosemond, Hanson & Zimmerman (2017) | USA | Understand how family decision-makers experienced goal-based decision making in advance of the death of their PLwD. | N=16<br><br>Daughter(n=13)<br>Spouse (n=1)<br>Cousin (n=1)<br>Brother (n=1) | Trusting relationship with nursing home staff. | Witnessing attentive care of their loved one.<br><br>Emotional support from staff (e.g., attending funeral of their loved one). | Reassure informal caregivers that goals of care are dynamic in dementia care.<br><br>Use lay terms when explaining processes and procedures.<br><br>Be proactive in organising conversations with informal caregivers after admission, to make them feel heard. | None discussed | Building trusting relationships with informal caregivers is key for effective 'goals of care' discussions.<br><br>Clarify that 'goals of care' refer to daily life as well as dying well. | None discussed |
| Ryan (2009) | UK | Conduct a literature review to understand the experiences of family caregiver decision-making about end-of-life care for their loved one living with dementia. | 10 articles | Accepting patient needs over family wishes to resolve tension and familial conflict.<br><br>Have knowledge and understanding of trajectory of their loved one's condition.<br><br>Collaboration and timely communication from professional care staff. | Educating themselves on their loved one's condition and understanding what the industry means by 'end-of-life' care. | Appreciate family caregivers as experts on their loved one living with dementia.<br><br>View family caregivers as equal in decision-making of PLwD care and come to a compromise which suits their wishes.<br><br>Inform families on treatment options before encouraging them to make decisions.<br><br>Collaborate and communicate in terms easily understood. | Limited to peer-reviewed literature from scientific journals. | Enhance relationships between family and professional care staff. | None discussed specifically. |
| Saini et al. (2016) | UK | Examine practices relating to end-of-life discussions with family members of PLwD residing in nursing homes and to explore strategies for improving practice. | Family caregivers (n=7)<br><br>Healthcare staff (n=19) | Receiving education on dementia prognosis and information on end-of-life care practices.<br><br>Having group discussions with care staff to discuss changing needs.<br><br>Having regular and timely discussion as disease progresses.<br><br>Trust in care staff.<br><br>Having feelings acknowledged by staff. | Regular discussions with care staff regarding end-of-life care, promotes capacity to consider options and make decisions.<br><br>Building trusting relationships with staff.<br><br>Reassurance from care staff.<br>.<br>Consider the situation as a dynamic process, and not make things permanent through over documentation. | Provide families with information before encouraging them to make advanced care plans and end-of-life care decisions.<br><br>Provide written information to support learning and document decisions.<br><br>Recognise and treat situation as fluid and offer numerous opportunities for discussions.<br><br>Provide time and space to have sensitive end-of-life discussions. | Volume of staff data outweighed volume of data from family caregivers.<br><br>Short interviews prevented in-depth conversations.<br><br>Short-term study | Designate a specific staff role to communicate and discuss end-of-life with family members and providing time and space to develop trusting relationships. | Dyad study educating professionals and families together and exploring level of understanding between the two. |

| Citation | Location | Research Questions/Aims | Sample (relationship to the PLwD) | Identified needs of informal caregivers | Coping mechanisms or support mentioned to improve own health and wellbeing | Preferences and/or suggestions for how informal caregivers could be involved in the decisions and care of PLwD | Limitations | Implications for practice | Avenues for future research |
|---|---|---|---|---|---|---|---|---|---|
| Sanders, Butcher, Swails & Power (2009) | USA | Explore how caregivers respond to the end stages of dementia with the assistance from hospice. | Family caregivers (N=27) | Consistent and thorough communication with the care setting.<br><br>Understanding the trajectory of their loved one's condition.<br><br>Education on what hospice and palliative care means. | Emotionally disconnecting with their loved one living with dementia, and reconnecting with aspects of their own lives which they felt they had 'lost'.<br><br>Preparation for death.<br><br>Limiting in-person interactions with their loved one living with dementia.<br><br>Conversations with other family and friends of the PLwD.<br><br>Avoid thoughts of "what could have been". | Being provided with sufficient information prior to admission, on the trajectory of their loved one's condition and set expectations, before making decisions.<br><br>Reassurance and support from the professional caregivers on the decisions informal caregivers make. | Limited sample according to ethnic and racial diversity. | Educate informal caregivers on end-of-life care, and trajectories of dementia. | Take accounts of experiences of informal caregivers in the earlier stages. |
| Sarabia-Cobo, Pérez, de Lorena, Nuñez & Domínguez (2016) | Spain | Describe the processes of decision-making used by families regarding treatments at the end-of-life of PLwD. | N= 84<br><br>Daughters (n=22)<br>Sons (n=9)<br>Spouses (n=12)<br>Siblings (n=16)<br>Nieces (n=11)<br>Grandchildren (n=5) | Accepting new role within the relationship with themselves and PLwD.<br><br>Good communication from care staff. | Focusing on care activities of daily living to maintain sense of control.<br><br>Care staff making time to answer questions and educate family on trajectory of disease.<br><br>Appreciating the little things and engaging in things you enjoy such as music and nature. | Make time to inform family member before asking them to make decisions on PLwD care.<br><br>Guidance from care staff on the outcomes of treatment options before making a decision.<br><br>Have a specific professional figure as a point of contact for decision discussions. | None discussed. | Ensure advanced care planning is done timely and sensitively, before crisis.<br><br>Educate and communicate the trajectory of dementia at end-of-life to families, to set expectations and eliminate uncertainty. | Examine interventions for advanced care planning. |
| Seiger, Cronfalk, Ternestedt & Norberg (2017) | Sweden | Explore how family members of PLwD describe their own experiences, before and after placing their relative in a nursing home. | N=10<br><br>Husband (n=1)<br>Wives (n=1)<br>Daughter (n=5)<br>Daughter-in-law (n=1)<br>Sons (n=2) | Care staff to confidently take majority of care responsibilities over and reassure families on admission.<br><br>The nursing home having a 'homelike' atmosphere and welcoming staff.<br><br>Knowledge and understanding of the complexity of dementia. | Delegate care responsibilities to care staff.<br>Regular informal chats with care staff to build rapport.<br>Balancing own life and health, alongside visiting their loved one.<br><br>Acknowledging and accepting their change in role as caregiver.<br><br>Using religion to justify the situation and decisions.<br><br>Acquainting the staff with their relative, making them view their loved one as a person. | Provide information on the trajectory of the disease, and information on how to interpret PLwD behaviours and their meanings before making care-based decision.<br><br>Value and respect family preferences and decisions, not pressuring or combatting them. | Small homogeneous sample within one care facility.<br><br>longer and repeated interviews combined with observations would have increased the trustworthiness. | Exercise family-centred care. | None discussed. |

| Citation | Location | Research Questions/Aims | Sample (relationship to the PLwD) | Identified needs of informal caregivers | Coping mechanisms or support mentioned to improve own health and wellbeing | Preferences and/or suggestions for how informal caregivers could be involved in the decisions and care of PLwD | Limitations | Implications for practice | Avenues for future research |
|---|---|---|---|---|---|---|---|---|---|
| Slape (2014) | Australia | Identify spiritual needs of family members during dementia and palliative care stages. | Family members (N=10) | Maintaining a meaningful relationship and staying connected to their loved one living with dementia.<br><br>Sense of trust with care staff and facility.<br><br>Seeing their loved one spiritually, emotionally, and emotionally content.<br><br>Confidence in ability to move on and think of the future. | Supportive relationships with care staff.<br><br>Maintaining relationships with extended family and friends for support.<br><br>Spiritual and emotional support being provided by facility for informal caregivers.<br><br>Finding meaning in the experience. | None discussed | None discussed | Providing or signposting family caregivers to readily available emotional or spiritual support. | None discussed. |
| Thompson, Hack, Rodger, John, Chochinov & McClement (2021) | Canada | Identify what information family caregivers of PLwD in nursing homes would deem useful in preparing them for end-of-life and assist them to make decisions about care. | Bereaved family caregivers (N=17) | Healthcare staff educating them on the type of dementia their loved one is diagnosed with.<br><br>Understanding dementia subtypes.<br><br>Receiving timely communication updating them on their loved one's condition and changes.<br><br>Meaningful interactions with PLwD. | Setting expectations on care and trajectory of their loved one's condition.<br>Maintaining personhood and sense of identity for loved one living with dementia.<br>Care staff normalising the experiences and emotions of family members, providing emotional support. | Providing regular opportunities to re-visit care-based decisions and amend with the dynamic trajectory of dementia.<br><br>Provide families with information before asking to make decisions.<br><br>Valuing family members as experts and treating them as respected member of the team. | Limited sample of four nursing homes in the same location.<br><br>Staff turnover was high in participants. | Get to know PLwD as a person, not just a resident.<br><br>Inform and educate families on what to expect, dementia itself, trajectory of disease early on. | Identify strategies that will engage PLwD in decision-making before their illness precludes them.<br><br>Identify strategies to optimise family member–health care provider communication. |
| Wladkowski (2016) | USA | Understand the experience of caregivers of PLwD, who experienced a 'live' discharge from hospice | N=24<br><br>Daughter (n=15)<br>Son (n=4)<br>Significant other (n=3)<br>Nonfamily appointed healthcare agents (n=2) | Preparation to lose the hospice care services.<br><br>Knowledge and understanding of dementia, and the unpredictability of its trajectory.<br><br>Understanding the impact on their grief, as caregivers. | Educating themselves on key terms such as 'end-of-life' care.<br><br>Support from professional care staff when having to face paperwork and lengthy documents. | Provide informal caregivers with sufficient information beforehand, so decisions can be made prior to a crisis. | Bias in participant selection from gatekeeper organisations, who may choose participants with positive and favourable experiences. | Training and education for care staff on the continuum of grief in this experience, so emotional and psychological support can be provided to informal caregivers throughout. | Further research into the grief process prior to and post-hospice. |
